# Seroprevalence of Anti-SARS-CoV-2 Antibodies in a Cohort of New York City Metro Blood Donors using Multiple SARS-CoV-2 Serological Assays: Implications for Controlling the Epidemic and “Reopening”

**DOI:** 10.1101/2020.11.06.20220087

**Authors:** Daniel K. Jin, Daniel. J. Nesbitt, Jenny Yang, Haidee Chen, Julie Horowitz, Marcus Jones, Rianna Vandergaast, Timothy Carey, Samantha Reiter, Stephen J Russell, Christos Kyratsous, Andrea Hooper, Jennifer Hamilton, Manuel Ferreira, Sarah Deng, Donna Straus, Aris Baras, Christopher D. Hillyer, Larry L. Luchsinger

**Affiliations:** Laboratory of Stem Cell Regenerative Research, Lindsley F. Kimball Research Institute, New York Blood Center, New York, NY 10065, USA; New York Blood Center Enterprises, New York, NY 10065, USA; Regeneron Genetics Center, Tarrytown, NY 10591, USA; Imanis Life Sciences, Rochester, MN 55901; Vyriad, Inc., Rochester, MN 55901; Mayo Clinic Department of Molecular Medicine, Rochester, MN 55905; Regeneron Pharmaceuticals, Inc., Tarrytown, NY 10591, USA

## Abstract

Projections of the stage of the Severe Acute Respiratory Syndrome-Coronavirus-2 (SARS-CoV-2) pandemic and local, regional and national public health policies designed to limit the spread of the epidemic as well as “reopen” cities and states, are best informed by serum neutralizing antibody titers measured by reproducible, high throughput, and statically credible antibody (Ab) assays. To date, a myriad of Ab tests, both available and authorized for emergency use by the FDA, has led to confusion rather than insight per se. The present study reports the results of a rapid, point-in-time 1,000-person cohort study using serial blood donors in the New York City metropolitan area (NYC) using multiple serological tests, including enzyme-linked immunosorbent assays (ELISAs) and high throughput serological assays (HTSAs). These were then tested and associated with assays for neutralizing Ab (NAb). Of the 1,000 NYC blood donor samples in late June and early July 2020, 12.1% and 10.9% were seropositive using the Ortho Total Ig and the Abbott IgG HTSA assays, respectively. These serological assays correlated with neutralization activity specific to SARS-CoV-2. The data reported herein suggest that seroconversion in this population occurred in approximately 1 in 8 blood donors from the beginning of the pandemic in NYC (considered March 1, 2020). These findings deviate with an earlier seroprevalence study in NYC showing 13.7% positivity. Collectively however, these data demonstrate that a low number of individuals have serologic evidence of infection during this “first wave” and suggest that the notion of “herd immunity” at rates of ∼60% or higher are not near. Furthermore, the data presented herein show that the nature of the Ab-based immunity is not invariably associated with the development of NAb. While the blood donor population may not mimic precisely the NYC population as a whole, rapid assessment of seroprevalence in this cohort and serial reassessment could aid public health decision making.

## Background

The Severe Acute Respiratory Syndrome Coronavirus (SARS-CoV)-2 pandemic has swept the global community with the United States reporting nearly 8.5 million confirmed cases and over 230,000 deaths from Coronavirus disease (COVID)-19.(1, 2) Transmission models of SARS-CoV-2, supported by studies of immune responses to related viral infections, suggest that recovery from infection could provide immunity to reinfection.(1, 3) Thus, the use of serological tests to identify those who have acquired antibodies (Abs) against SARS-CoV-2 (seroconversion) and the frequency of seroconversion in the population (seroprevalence) is a powerful means with which to guide public health policies.(4, 5) The term ‘hotspots’ has emerged to describe regions of high infectivity that appear and then recede as the pandemic evolves. It is important to ascertain the frequency of SARS-CoV-2 seropositivity in regional populations to estimate the risk of infection associated with newly developing or receding COVID-19 hotspots.

As natural infection continues to persist, and vaccine distribution commences, serologic assays will be vital in monitoring the development of herd immunity, also called community or population immunity, which refers to the point at which enough people are sufficiently “protected”, and person-to-person transmission is unlikely. Reaching this milestone will, in effect, herald the end of the COVID-19 pandemic. Therefore, population-wide serological assessment and reassessment are critical, and the tests employed need to be reliable, credible, reproducible and high throughput. Furthermore, it is important to understand the degree of correlation of any given assay’s “reactivity” with the presence of neutralizing antibody (Nab). These data, then, can be used to assist public health officials in modeling projections and in informing policy making decisions including the safe “reopening” of cities, states, and regions.

The performance and sensitivity of COVID-19 serology assays is myriad in platform (lateral flow, ELISA, etc.) and variable in terms of sensitivity and specificity.(6, 7) Such assays rely on detection and quantification of antibodies that recognize specific SARS-CoV-2 antigens including the four major structural proteins; spike (S) protein (containing the S1 domain and RBD motif), nucleocapsid (NP) protein, membrane (M) protein, and envelop (E) protein.(8) Research conducted on 2005 SARS-CoV-1 and Middle East respiratory syndrome Coronavirus (MERS-CoV), which are highly related to SARS-CoV-2, found that recovered individuals produced the strongest immunogenic antibodies against antigens of the S and N proteins.(9) Thus, the development of serological tests for SARS-CoV-2 antibodies has focused heavily on the detection of antibodies against these viral proteins.

As above, antibody-based tests vary considerably in both technology (platform) and target antigen (design) which led to, in May 2020, the FDA reversing its emergency use authorization (EUA) and approval policies in order to help ensure that reliable tests could be used to accurately measure seroconversion in populations. Some tests have received emergency use authorization but population-wide data are limited, and continuous monitoring is necessary to be of practical importance.

Variability in test characteristics, particularly sensitivity, implies that there may not yet be an *ideal* test design and instrument platform, which can lead to variability and potential bias in the estimation of the level of immunity in various locales or subpopulations.(10, 11) However, two platforms have been widely cited: 1) in-house enzyme linked immunosorbent assays (ELISA), and 2) high-throughput serological assays (HTSA). ELISAs offer wide flexibility for research laboratories to select virtually any antigenic protein of interest and assay patient sera to provide highly sensitive, quantitative results. HTSAs are more suitable to clinical laboratories processing large volumes of samples. Although HTSAs offer a narrower selection of antigen choices, these platforms offer high-throughput capacity, high sensitivity and can be integrated into clinical lab testing facilities. The resulting expectation of antibody development is an association with antiviral activity and acquisition of immunity against future viral infection. However, only a subset of virus-specific antibodies will be neutralizing, and the levels of SARS-CoV-2-specific neutralizing antibodies necessary to confer protective immunity following infection or vaccination across the population are not known. Thus, studies that evaluate serological test designs are necessary to associate a serological result with a probability of immunity.

New York City (NYC) was one of the first epicenters of the COVID-19 pandemic and possesses the highest case count per capita of anywhere in the United States.(12) Seroconversion, therefore, is likely to be substantial in a random sampling of NYC residents. Moreover, the true number of COVID-19 cases may be underreported, resulting in inaccurate case estimates (incidence) and morbidity and mortality rates of SARS-CoV-2.(13)

The objectives of the study reported herein were to determine the seroprevalence of anti-SARS-CoV-2 Ab in blood donors in the NYC metro area at a specific point in time four months after the first NY case, as a surrogate for the population as a whole, an indicator of the stage of the epidemic, and as a baseline for future reassessments, using commercially available serology tests, and characterize the Ab responses in ELISAs and a neutralizing antibody assays, allowing us to ultimately inform city, state and nation-wide efforts to mitigate the pandemic and its attendant social and economic strife.

## Results

### Characteristics of the NYC Blood Donor Population

To estimate seroprevalence, 1,000 blood donor plasma samples were randomly collected from NYBC blood donation centers between June and July 2020, encompassing regions proximal to NYC, including Long Island, Westchester County and New Jersey (**Figure 1A**). To characterize donors demographically, we cross-referenced donor data to the 2010 U.S. Census dataset.(14) Donors ranged in age from 16 to 78 years with a median age of 48 years (95% CI: 46-49 years), which was older than the New York City median age of 35.5 years and deviated from a Gaussian distribution (**Figure 1B**, r^2^=0.708). The donor group also included significantly fewer female donors (38.5%) compared to 52.5% citywide (**Figure 1C**). Donors that did not respond to ethnicity or reported as ‘Other’ composed 15.6% of the donors. Among donors that responded, the distribution of donor race/ethnicity was 73% white, 3.6% black, 3.4% multi-race, and 4.4% Asian, compared to an average NYC Metro distribution of 44% white, 25.55% black, 3.99% multi-race, and 12.7% Asian (**Figure 1D**). These data show skewing of blood donors from the NYC demographics in many categories, which is a known characteristic of the blood donor population.

**Figure 1:**
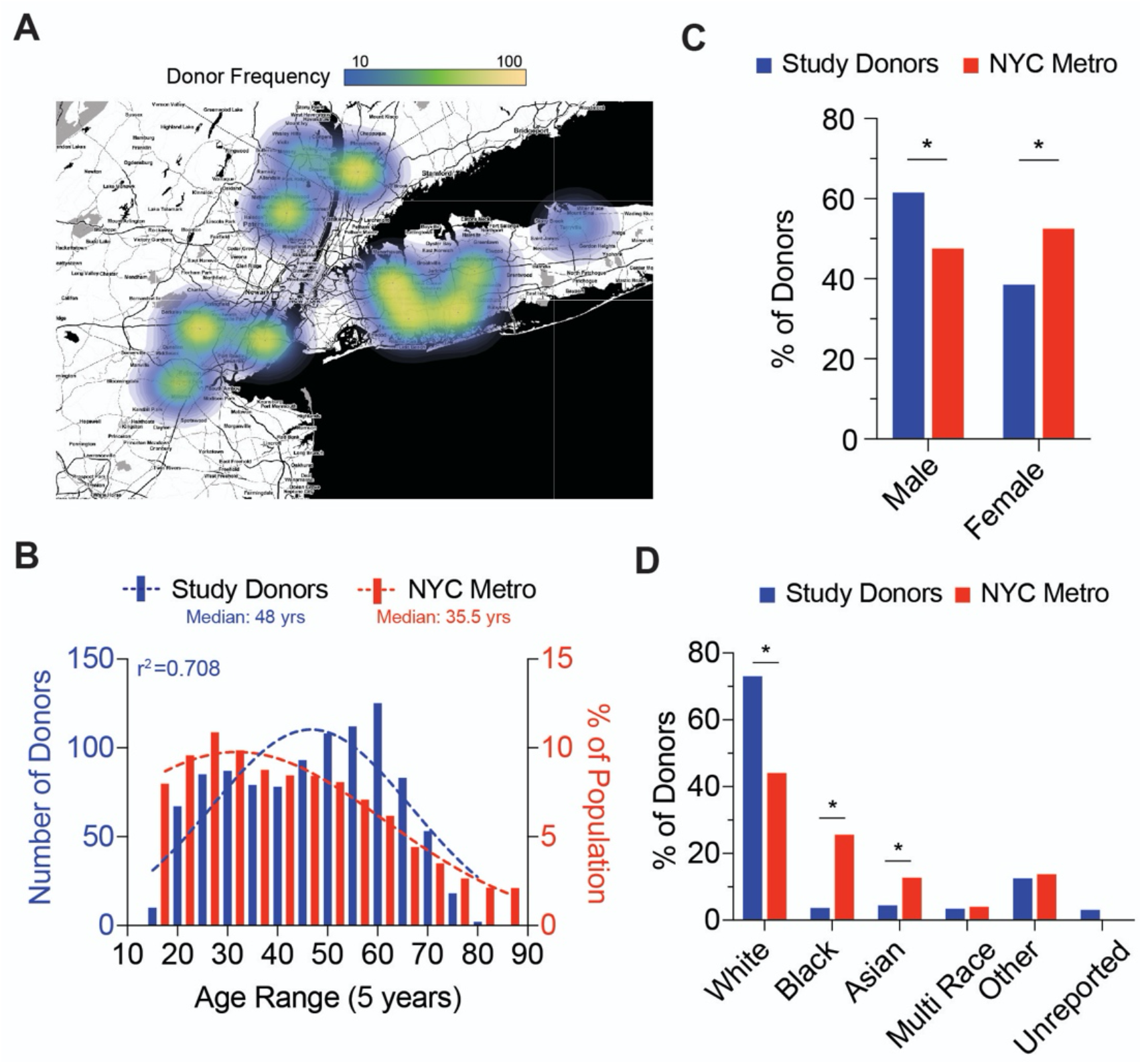
Blood Donor Demographics of NYC Metro Area. **A;** Choropleth of donation site locations used for collection of blood donor samples. Heatmap (gradient bar, top) indicates frequency of donors collected per site. **B;** Distribution of NYC Metro area donor age (red bars) compared to NYC demographics (blue bars). Dotted lines represent best fit to a Gaussian distribution and r2 value represents calculated goodness of fit to distribution plot. **C;** Gender frequency of NYC Metro area donors (red bars) compared to NYC demographics (blue bars). Chi-square test for goodness of fit to observed (donors) versus expected (NYC demographics) results; *p<0.01 **D;** Ethnicity frequency of NYC Metro area donors (red bars) compared to NYC demographics (blue bars). Chi-square test for goodness of fit to observed (donors) versus expected (NYC demographics) results; *p<0.01

### High Throughput Serological Estimates

To quantify SARS-CoV-2 seroprevalence in donor samples, the Ortho Clinical Diagnostics VITROS Total Ig Test (Ortho) and the Abbott Labs Architect SARS-CoV-2 IgG (Abbott) HTSA assays were used. Results of the Ortho test yielded 121 positive donors while the Abbott test showed 109 positive donors (**Figure 2A**). Adjusting for sample size effect, the estimated seroprevalence rate using the Ortho HTSA was 12.1% (95% CI: 10.2 – 14.27%) while the Abbott test indicates a seroprevalence rate was 10.9% (95% CI: 9.1% - 12.9%). In total, 128 donors were seropositive by either HTSA test, with 102 donors (79.69%) testing positive for anti-SARS-CoV-2 antibodies using both the Ortho and Abbott tests, and 19 (14.84%) or 7 (5.47%) of donors testing positive using only the Ortho or Abbott test, respectively (**Figure 2B**). The median results using the Ortho test for seropositive donors was 414 (n=121, 95% CI: 320.0-466.0, IQR: 135.0-692.5), representing a 5,900-fold increase over the median Ortho result for seronegative donors which was 0.07 (n=879, 95% CI: 0.07-0.07, IQR 0.05-0.10) (**Figure 2C**). The median Abbott test result for seropositive donors was 4.1 (n=109, 95% CI: 3.56-4.77, IQR: 2.77-5.915), representing a 130-fold increase over the median Abbott result for the seronegative donors which was 0.03 (n=891, 95%CI: 0.02-0.03, IQR: 0.02-0.5) (**Figure 2D**).

**Figure 2:**
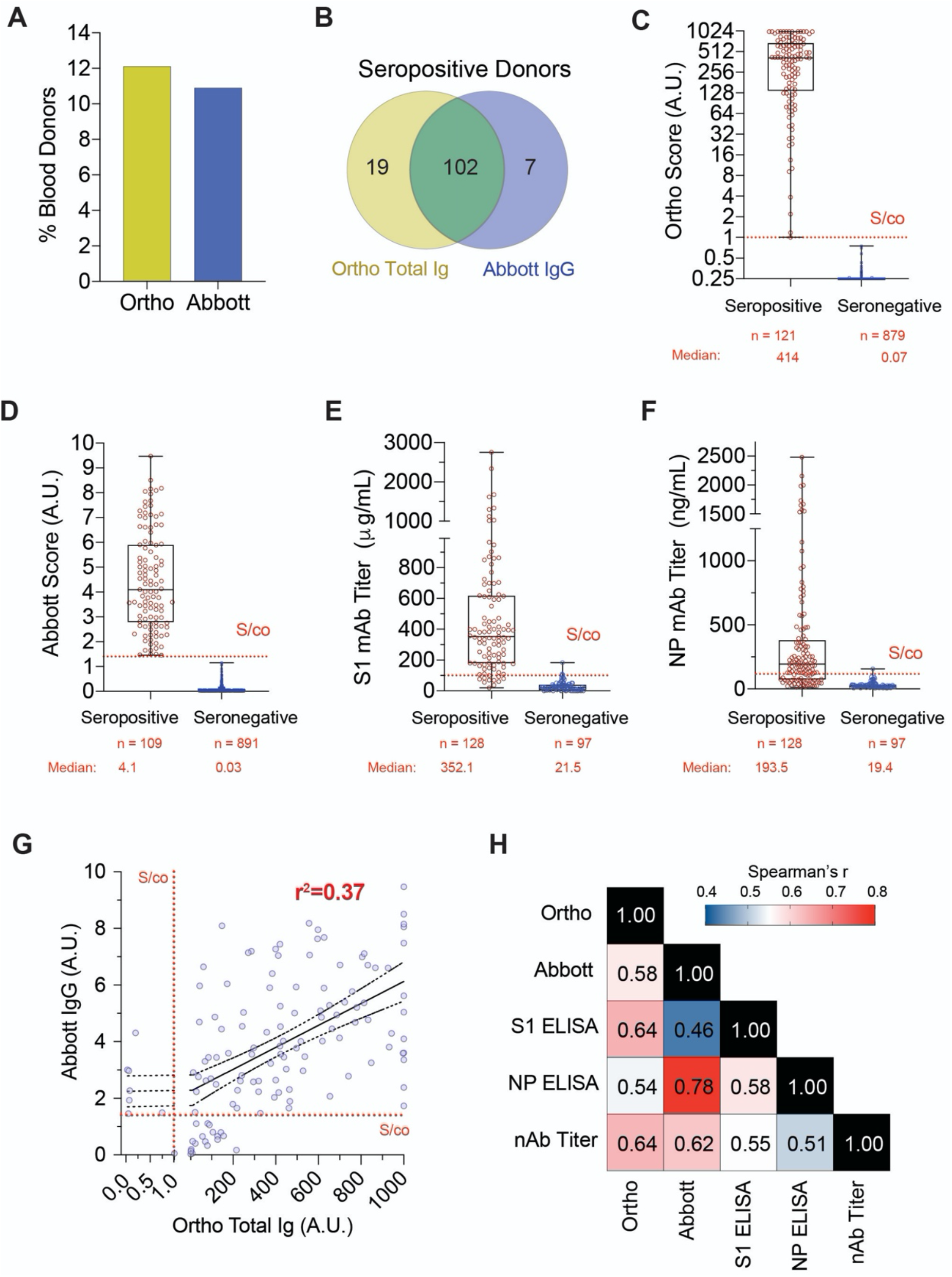
Serological and Neutralizing Activity Analysis of NYC Metro Blood Donors. **A;** Frequency of NYC Metro area seropositive donors as determined using the Ortho Total Ig (yellow bar) or Abbott IgG (blue bar) HTSA assays. **B;** Venn diagram of donors determined to be seropositive using the Ortho (yellow) or Abbott (blue) HTSA assays. Seropositive donors that were reactive for both tests are indicated in overlap (green). **C;** Distribution of Ortho HTSA serological results between seropositive (red dots) and seronegative (blue dots) as determined by the Ortho HTSA assay. Median value and sample number is shown below graph. Dotted line shows S/co value (1.00 A.U.). **D;** Distribution of Abbott HTSA serological results between seropositive (red dots) and seronegative (blue dots) as determined by the Abbott HTSA assay. Median value and sample number is shown below graph. Dotted line shows S/co value (1.4 A.U.) **E;** Distribution of S1 ELISA serological results between seropositive (red dots) and seronegative (blue dots) as determined by either HTSA assay. Median value and sample number is shown below graph. Dotted line shows S/co value (100μg/mL). **F;** Distribution of NP ELISA serological results between seropositive (red dots) and seronegative (blue dots) as determined by either HTSA assay. Median value and sample number is shown below graph. Dotted line shows S/co value (100 μg/mL). **G;** Linear regression of seropositive donor of HTSA results. Dotted lines denote signal to cutoff (S/co) for each test and goodness of fit, r^2^, is shown. **H;** Spearman correlation coefficients, r, between each serological assay.

The gold-standard of serological quantification is the ELISA assay. To compare HTSA results using our in-house SARS-CoV-2 ELISA assays, we analyzed donor plasma for antibodies using S1 and NP antigens. Using the S1 ELISA (**Figure 2E**), the median value for seropositive donors was 352.1 μg/mL (n=128, 95% CI: 312.0-399.8 μg/mL, IQR: 179.9 – 617.2 μg/mL) and the median value for negative donors was 21.5 μg/mL (n=97, 95% CI: 17.32 – 26.77 μg/mL, IQR: 6.29 – 38.79 μg/mL). Using the NP ELISA (**Figure 2F**), the median value for seropositive donors was 193.5 ng/mL (n=128, 95% CI: 155.6 ng/mL-226.7ng/mL, IQR: 74.00 ug/mL-380 ug/mL) and the median value for negative donors was 19.38 ng/mL (n=97, 95% CI: 15.74 ng/mL - 24.20 ng/mL, IQR: 12.79 ug/mL - 31.49ug/mL). Interestingly, seropositive donors for Ortho and Abbott tests showed 88.2% and 84.5% above the S/co value for the S1 ELISA assay. Expectedly, seropositive donors negative by S1 ELISA assays had relatively low HTSA scores (data not shown), which suggests HTSA assays have higher sensitivity than traditional ELISA methodology. Linear regression of Ortho and Abbott tests (**Figure 2G**) showed a modest goodness-of-fit (r^2^=0.37) indicating that while HTSA test scores are positively associated, a high degree of variation within donors exists between HTSA test results. Taken together, these data confirm that a wide range of serological results are prevalent in the NYC metro population and HTSA platforms have the highest sensitivity to quantify serological results with which to estimate seroprevalence.

### Neutralizing Activity of NYC Blood Donors

Antiviral antibodies can inhibit viral particles from infecting target cells and constitute an important form of immunity to future viral exposure; particularly in relation to effective vaccination. In the case of SARS-CoV-2, such assays require biosafety level 3 (BSL-3) facilities and highly trained personnel. To overcome this limitation and expedite testing, we employed a ‘surrogate virus’ neutralization assay to quantify nAb levels present in donor plasma, which differs from conventional SARS-CoV-2 pseudovirus particles in that surrogate virus retains replication potential and is thus more analogous to live SARS-CoV-2. The results of the neutralization end point titer (NT_100_) assays are summarized in **Table 1**. The majority (87.4%, n=90) of Ortho seropositive donors (n=121) were positive for nAbs, while 18 samples (14.9%) had indeterminant levels of nAbs and 13 samples (12.6%) were negative for neutralizing activity. Ortho seronegative donors (n=104) showed 1 positive (0.9%) and 2 (1.8%) indeterminant samples for neutralizing activity. The majority (92.4%, n=86) of Abbott seropositive donors (n=109) were also positive for nAbs, with 16 samples (14.7%) having indeterminant levels of nAbs and 7 samples (7.6%) being negative for neutralizing activity. Abbott seronegative donors (n=116) showed 5 positive (4.3%) and 4 (3.4%) indeterminant samples for neutralizing activity. It was noted that all samples positive for neutralization activity were positive for at least one HSTA assay, while 14.9% of seropositive samples were negative for neutralization activity (**Table 2**). These data illustrate the that serological assays, particularly those with values near the S/co value for each assay, may not reliably correspond to bona fide neutralization activity.

**Table 1:** Correlation of Serological Results with Neutralization Activity.

| Neutralization Result |  |  |  |  |
| --- | --- | --- | --- | --- |
| Ortho HTSA | Positive | Negative | Indeterminate/<br>Borderline | Total |
| Seropositive | 90 | 13 | 18 | 121 |
| Seronegative | 1 | 89 | 2 | 104 |
|  |  |  |  | 225 |
| Abbott HTSA | Positive | Negative | Indeterminate/<br>Borderline | Total |
| Seropositive | 86 | 7 | 16 | 109 |
| Seronegative | 5 | 95 | 4 | 116 |
|  |  |  |  | 225 |

**Table 2:** Neutralization Activity as a Function of Serological Results.

| HTSA Result | Neutralization Result |  |  |
| --- | --- | --- | --- |
|  | Positive | Indeterminate/<br>Borderline | Negative |
| Ortho Only | 5 | 4 | 10 |
| Abbott Only | 1 | 2 | 4 |
| Double Positive | 85 | 14 | 3 |
| Double Negative | 0 | 0 | 97 |
| Total Samples Tested | 91 | 20 | 114 |
| Percent HTSA Reactive | 100% | 100% | 14.9% |

The semi-quantitative NT_100_ method showed that titers for seropositive samples varied between donors. Reciprocal dilution factor values ranged from <80 to 1,280 (**Figure 3A**). Analysis of the 128 seropositive samples revealed 21.9% were below the LOD at the 1:80 dilution (the lowest dilution tested in this analyses) and 7.0% were considered ‘indeterminant’, due to suspected sample interference. We found low NT_100_ titers of 80 and 160 comprised 30.5% and 29.7% of BD samples, respectively, constituting over half of seropositive blood donors. Moderate NT_100_ titers of 320 and 640 accounted for 5.5% and 4.7% of donors while the highest nAb titers of ≥1280 described 0.8% of seropositive samples. These data indicate that, similar to serology results, nAb levels against SARS-CoV-2 are highly variable and are skewed towards low neutralizing activity within seropositive blood donors in the NYC metro area.

**Figure 3:**
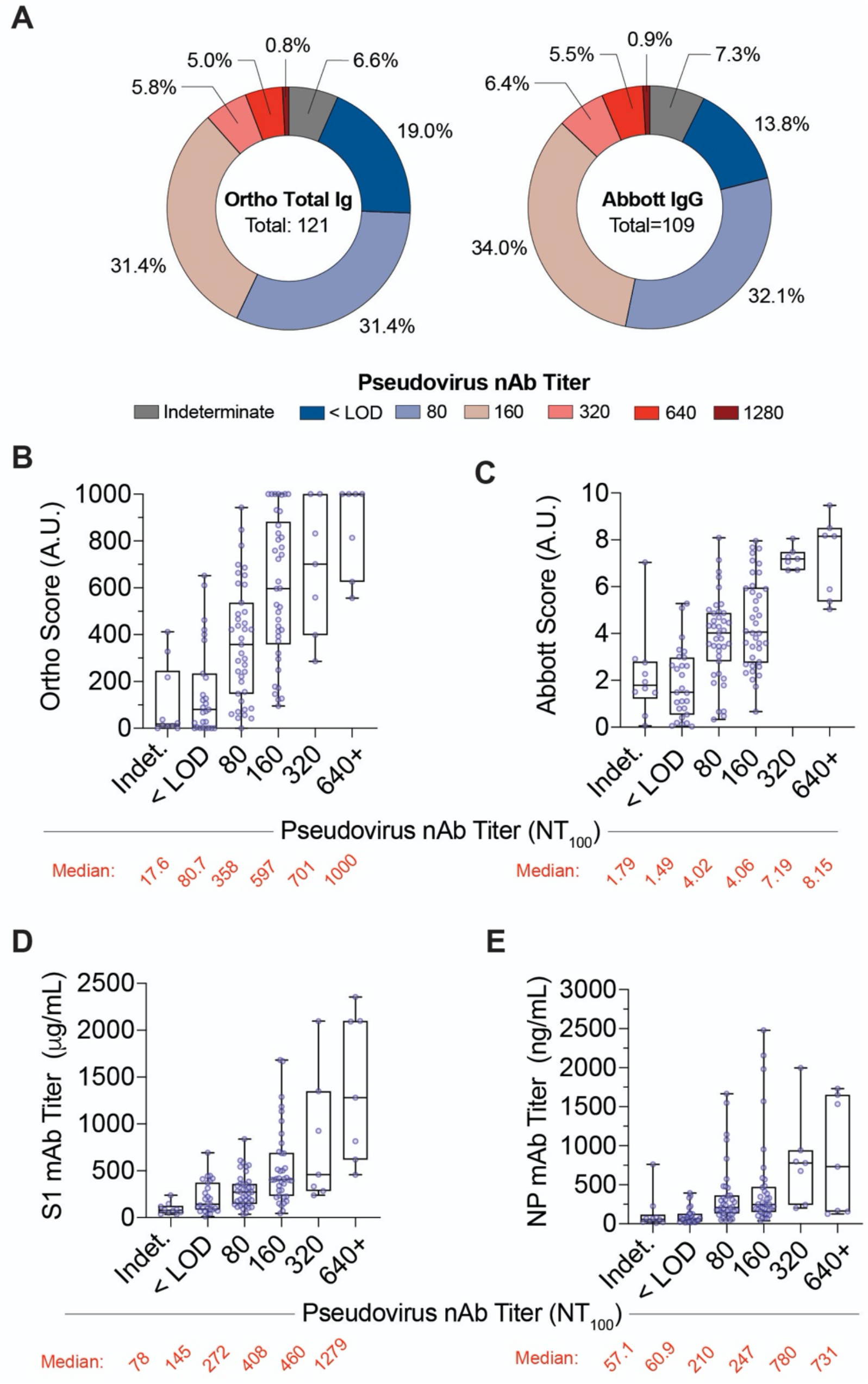
Correlation of NYC Metro Donor Serological Results with Neutralization Activity. **A;** Frequency of Ortho HTSA (left) or Abbott HTSA (right) seropositive donor pseudovirus neutralization end-point titers. **B-E;** Box plots of seropositive donor serology results using the Ortho HTSA, Abbott HTSA, S1 ELISA and NP ELISA for each category of neutralization end point titers. Boxes and whiskers denote 1^st^ and 3^rd^ quartiles and range, respectively. Median serology value of each category is shown below graph.

It remains infeasible to implement neutralization assays as a measurement of antiviral antibodies at the scale of the general population. While many serology tests have been developed, evidence as to the predictive value between SARS-CoV-2 serology test results and neutralizing activity continues to be an important validation for the medical and scientific community. To this end, we examined the correlation between serology and neutralization assays in the CP donor samples. Spearman’s correlation analysis of all five serological analyses showed a high degree of positive association between all tests (**Figure 2H**). The Ortho and Abbott HTSA tests showed the highest degree of correlation with neutralization activity (Ortho: r= 0.64, Abbott: r=0.62), followed by the S1 (r=0.55) and NP (r=0.51). Interestingly, the Ortho HTSA and S1 ELISA, and Ortho HTSA and nAb assay showed the highest correlation (r=0.64), further validating the SARS-CoV-2 spike S1 protein as an important target of neutralizing function. These data confirm that established HTSA and ELISA assays strongly correlate with neutralizing activity. As expected, the Ortho test, which measures anti-spike antibodies, showed a higher degree of correlation with the S1 ELISA titers (r=0.639) while the Abbot test, which measures anti-NP antibodies, showed a high degree of correlation with the NP ELISA titer (r=0.778).

Further, median values for both HTSA assays increased with higher neutralizing assay titers (**Figure 3B, 3C**) and this observation was also observed in ELISA assays (**Figure 3D, 3E**). These data highlight the utility of HTSA and ELISA assays to predict neutralization activity of plasma samples.

Similarly, we evaluated the correlation between donor-reported SARS-CoV-2 polymerase chain reaction (PCR) testing, SARS-CoV-2 serology, and neutralization. 273 donors (27.3%) reported having had a PCR test, with 44 (16.1%) reporting to have had a positive PCR result, 203 reporting to have had a negative PCR result, and 750 donors who were either not tested or were awaiting PCR results (**Table 3**), though date of PCR test was not available. Of the 44 donors reporting to have had a positive PCR result, 43 (97.7%) were seropositive, with 39 (88.6%) being both Ortho seropositive and Abbott seropositive, and an additional 3 being Ortho seropositive, and 1 being Abbott seropositive. Donors reporting to have had a positive PCR result were also tested for neuralization activity, with 38/44 (86.4%) samples positive, 5 indeterminate, and 1 negative for neutralizing activity (**Table 4**). Though the amount of time between positive SARS-CoV-2 PCR test result and blood donation is not known, CP eligibility requires that the donor be 14 days from the date of last symptom. Whether or not indeterminate/negatives may have seroconverted subsequent to the donation analyzed, these results show strong correlation between positive SARS-CoV-2 PCR test results, seropositivity, and neutralization activity and may be suggestive of longitudinal immunity.

**Table 3:** Correlation of Serological Results with Self-Reported SARS-CoV-2 PCR Results.

| Self-Reported SARS-CoV-2 PCR Test Result | Seropositive Donors by HTSA |  |  | Seronegative Donors | Total |
| --- | --- | --- | --- | --- | --- |
|  | Ortho/Abbott | Ortho | Abbott |  |  |
| Positive | 39 | 3 | 1 | 1 | 44 |
| Negative | 16 | 5 | 1 | 181 | 203 |
| Untested/No result | 47 | 11 | 5 | 690 | 750 |

**Table 4:** Correlation of Neutralization Results with Self-Reported SARS-CoV-2 PCR Results.

| Neutralization Result |  |  |  |  | 373 |
| --- | --- | --- | --- | --- | --- |
| SARS-CoV-2 PCR Test Result | Positive | Negative | Indeterminate | Total |  |
| Positive | 38 | 1 | 5 | 44 | 374 |
| Negative | 14 | 27 | 2 | 43 |  |
| Untested/No result | 37 | 90 | 11 | 138 |  |

## Discussion

COVID-19 antibody testing has entered public discourse as an important metric in monitoring the evolution of the SARS-CoV-2 outbreak. Ultimately, the application of antibody testing could be clinically informative as to the degree of immunity incurred by recovered patients or vaccinated individuals. Random blood donor screening is a practice that is readily feasible using blood banking infrastructure to rapidly screen regional populations for seroprevalence monitoring. This is the first study to evaluate a large cohort of random blood donors in the NYC metro area for SARS-CoV-2 antibodies. However, we recognize the limitations of the current study include a lack of generalizability as a consequence of the modestly skewed demographics of blood donors and the general population as a whole, and that this may impact the conclusions of the results. In fact, seroprevalence has been suggested to be higher in specific racial/ethnic communities based on recent studies.(15) Thus, more inclusive and complete seroprevalence studies will need to be performed in the future. Nonetheless, the authors believe that using blood donor in serial studies is an important, if indirect, measure of community immunity.

In this study, we found the Ortho Total Ig and Abbott IgG HTSA assays estimate a ∼10.9-12.2% SARS-CoV-2 seroprevalence in July of 2020 in the NYC Metro area. Moreover, we found that ELISA assays, which are the gold-standard of serological quantification, corresponded with seropositive classification of donors as detected by HTSAs, thus validating the use of ELISAs in population studies. Further, in seropositive blood donors we observed a wide range of anti-SARS-CoV-2-neutralizing activity that was skewed towards low to moderate NT_100_ titers. This trend is in agreement with our previous investigation of convalescent plasma donors(16) and a study of patients recovering from COVID-19, both of which also showed large variability and modest levels of neutralizing activity in plasma units.(17)

Our estimation of the NYC Metro area blood donor seroconversion is in agreement with other reports from state and local departments of health. Seroconversion in a study of Bergen County, NJ was estimated to be 12.2% in June of 2020.(18) Seroconversion among hospital workers in New York City was estimated to be 13.7% as of June of 2020.(19) The overall seroprevalence in New York City, at the peak of the epidemic, was estimated to be 21% with some communities as high as 68% using data from emergency care clinics.(20) This is juxtaposed to neighboring states, such as Rhode Island, where we estimated seroconversion to be 0.6% among blood donors in May 2020.(21) Given the early introduction of SARS-CoV-2 in the NYC Metro area in March of 2020 as an initial, and possibly largest, ‘hot-spot’ in the United States, the estimated seroprevalence in this study may be lower than anticipated due to naturally waning antibody titers(22) (or due to demographics of donor population relative to the NYC population.

## Conclusion

In conclusion, we estimate the seroprevalence of NYC metro blood donors to be approximately 1 in 8 donors during the month of July 2020 and four months post the commencement of the epidemic in NY. While it is slightly lower than another study using a NYC population of healthcare workers during a similar time period, who, in all likelihood, had higher than typical exposure rates,(19) our findings demonstrate a comparable seroprevalence estimate can be discerned using a widely accessible blood donor population and it an important metric during this catastrophic outbreak. This strategy can therefore be leveraged in its design for future studies to implement rapid seroconversion/seroprevalence monitoring. Furthermore, considering the possibility that this may be an underestimate of the metropolitan population, these conclusions suggest that in the absence of a vaccine, “background” or “herd” immunity continues to be low at four months post-commencement, and, now eight months into the US pandemic, it is probable that the susceptible population remains very high, and possibly at ∼80% or greater.

## Methods

### Whole Blood Donors and Sample Preparation

From June 16, 2020 – July 15, 2020, consecutive NYC metro donors (n=1,000) received a 2-question survey, provided demographic information and completed a blood donation. Choropleth was generated in house from donor zip code prefix data using the web tool, http://www.heatmapper.ca/geocoordinate/. Plasma was isolated from whole blood samples collected in citrate tubes. Samples were extracted, aliquoted to minimize freeze-thaw cycles, and stored at -80°C. Donor blood samples were tested using the Ortho VITROS™ SARS-CoV-2 Total Ig assay, Abbott SARS-CoV-2 IgG assay, in-house ELISAs, and the Vyriad IMMUNO-COV™ neutralization assay as described with some modifications.(23) The IMMUNO-COV assay performed here differed from that which was described in the referenced publication in that: 1) plasma samples were heat-inactivated instead of serum samples, which is necessary due to thermal coagulation and 2) neutralization activity was quantified using neutralization end point titer (NT_100_) method and not a standard curve.

### High-throughput Serology Assays

Plasma samples were barcoded and dispatched to Rhode Island Blood Center (RIBC). Samples were analyzed using the Abbott SARS-CoV-2 IgG chemiluminescent microparticle immunoassay using the Abbott Architect i2000SR (Abbott Core Laboratories), as well as the VITROS Immunodiagnostic Products Anti-SARS-CoV-2 Total Test using the VITROS 5600 (Ortho Clinical Diagnostics). All assays were performed by trained RIBC employees according to the respective manufacturer standard procedures.

### Virus Neutralization Assays

Plasma samples were heat-inactivated for 30 min at 56°, then clarified by centrifugation for 5 min. at 12,000 x g and assayed using a surrogate virus SARS-CoV-2 neutralization assay. A modified version of the IMMUNO-COV™ assay (19), was used in which each plasma sample was serially diluted and assayed at a total of six dilutions, starting at 1:80. The virus neutralizing titer was determined as the reciprocal of the highest dilution at which the sample was still positive for neutralization based on assay performance relative to a pre-defined calibrator consisting of monoclonal anti-spike antibody.

### In-house SARS-Cov2 Binding-Antibody ELISAs

Flat-well, nickel-coated 96 well ELISA plates (Thermo Scientific; USA) were coated with 2 ug/mL of recombinant S1 spike protein, nucleocapsid protein, or Receptor Binding Domain (RBD) spike protein specific to SARS-CoV-2 in resuspension buffer (1% Human Serum Albumin in 0.01% TBST) and incubated in a stationary humidified chamber overnight at 4°C. On the day of the assay, plates were blocked for 30 min with ELISA blocking buffer (3% W/V non-fat milk in TBST). Standard curves for both S1 and RBD assays were generated by using mouse anti-SARS-CoV-2 spike protein monoclonal antibody (clone [3A2], ABIN2452119, Antibodies-Online) as the standard. Anti-SARS-CoV-2 Nucleocapsid mouse monoclonal antibody (clone [7E1B], bsm-41414M, Bioss Antibodies) was used as a standard for nucleocapsid binding assays. Monoclonal antibody standard curves and serial dilutions of donor sera were prepared in assay buffer (1% W/V non-fat milk in TBST) and added to blocked plates in technical duplicate for 1 hr with orbital shaking at room temperature. Plates were then washed three times with TBST and incubated for 1 hr with ELISA assay buffer containing Goat anti-Human IgA, IgG, IgM (Heavy & Light Chain) Antibody-HRP (Cat. No. ABIN100792, Antibodies-Online) and Goat anti-Mouse IgG2b (Heavy Chain) Antibody-HRP (Cat. No. ABIN376251, Antibodies-Online) at 1:30000 and 1:3000 dilutions, respectively. Plates were then washed three times, developed with Pierce TMB substrate (Thermo Scientific; USA) for approximately 5-7.5 min, and quenched with 3 M HCl. Absorbance readings were collected at 450 nm. Standard curves were constructed in Prism 8.4 (Graphpad Software Inc.) using a Sigmoidal 4PL Non-Linear Regression (curve fit) model.

### Estimated Seroprevalence & Statistical Calculations

For HTSA assays, seroprevalence was estimated using the Wilson Bayesian statistical method.(24) Data and statistical analyses were performed and presented using Prism 8.

## Data Availability

All raw data will be made available upon request to the corresponding author.

## Authors’ Contributions

DKJ, DJN, HC and JY performed serology experiments and analyzed donor demographic analyses. JH coordinated neutralization assays. MJ and DS managed the collection and distribution of donor samples. LLL managed the collection of data, figures and statistical analyses. LLL, JH, AB and CDH co-wrote the manuscript.

### Acknowledgements

We thank Jill Alberigo, Amanda Brites and Kelly Brightman from Rhode Island Blood Center for their help with performing the Ortho Anti-SARS-CoV-2 Total Test and the Abbott SARS-CoV-2 IgG test.

